# DNA Methylation Age Acceleration, Type 2 Diabetes, and its Complications: Cross-sectional and Longitudinal Data from the Berlin Aging Study II (BASE-II)

**DOI:** 10.1101/2022.06.28.22276991

**Authors:** Valentin Max Vetter, Johanne Spieker, Yasmine Sommerer, Nikolaus Buchmann, Christian Humberto Kalies, Vera Regitz-Zagrosek, Lars Bertram, Ilja Demuth

**Author notes:** **Corresponding author:** Ilja Demuth (Ph.D.), Charité - Universitätsmedizin Berlin, Lipid Clinic at the Interdisciplinary Metabolism Center, Biology of Aging Group, Augustenburger Platz 1, 13353 Berlin.

## Abstract

Patients with diabetes mellitus are at risk for micro- and macrovascular complications that are responsible for a substantial part of the individual health burden and socio-economic costs. Therefore, implementable risk scores are needed to improve targeted prevention for patients that are particularly susceptible to complications. The “epigenetic clock” estimates an individual’s biological age using DNA methylation profiles and was previously shown to be associated with morbidity and mortality.

In this study, we examine older adults of the BASE-II study that were reexamined on average 7.4 years after baseline assessment as part of the GendAge study. For DNA methylation age (DNAmA) estimation we used the 7-CpG clock which was available for two timepoints (n=1,071 at follow-up). In addition, we determined epigenetic age using Horvath’s clock, Hannum’s clock, PhenoAge and GrimAge which were available at follow-up only (n=1,067). The deviation of DNAmA from chronological age, DNA methylation age acceleration (DNAmAA), was calculated as residuals of a leukocyte cell count adjusted linear regression analysis. Diabetes associated complications were assessed with the Diabetes Complications Severity Index (DCSI).

Cross-sectionally, a statistically significant association between oral glucose tolerance test results and Hannum (ß=0.8, SE=0.3, p=0.02, n=762) and PhenoAge DNAmAA (ß=0.8, SE=0.3, p=0.003, n=762) was found. PhenoAge was also associated with fasting glucose (ß=0.3, SE=0.1, p=0.013, n=966). In contrast, we observed no cross-sectional association after covariate adjustment between DNAmAA and a diagnosis of diabetes mellitus with any of the five clocks employed. This was true for longitudinal analyses with the 7-CpG clock as well. However, longitudinal analyses showed that every year in the 7-CpG-based DNAmAA estimate at baseline increased the risk for developing of one or more additional complications or worsening of an already existing complication during the follow-up period by 11% in male participants with diabetes mellitus type 2. This association persisted after adjustment for DCSI at baseline, chronological age, smoking, alcohol, diabetes medication, and BMI (OR =1.11, p=0.045, n=56). No statistically significant association was found in the subgroup of women or when the whole dataset was analyzed (p>0.05).

Although our findings still need to be independently validated, the 7-CpG clock appears to be a promising biomarker which is informative about the individual risk for diabetic complications independent of age, sex, lifestyle factors, or complications at baseline.

## Introduction

The number of people diagnosed with diabetes mellitus has increased fivefold to 537 million over the last 31 years [1, 2] and is projected to increase further to 784 million until 2045 [1]. The disease’s impact on well-being and daily living differs between individuals and is substantially driven by its complications [3]. Besides acute complications like diabetic ketoacidosis, hyperosmolar coma, and hypoglycemia, chronic microvascular (retinopathy, nephropathy and neuropathy) and macrovascular (coronary artery disease, peripheral artery disease and stroke) complications [4] can result from diabetes mellitus. While some of these complications are life-threatening, others are at least disabling and can result in substantial emotional and financial burden for the affected individual and their family members [2, 5]. Furthermore, the resulting costs for the health care system are high [1] and expected to rise further [6]. A substantial part of these costs was attributed to diabetic complications [2, 7-10]. Although complications cannot be prevented completely by the currently available therapeutic options, early interventions can delay their onset [11] and prevent potentially severe disease progression. Although several risk scores for diabetic complications exist (reviewed in [12-18]), they are not used regularly in clinical practice [19] or are recommended by health organizations [2, 19, 20]. Therefore, the demand for a risk score with high predictive power that is based on reliable and well-assessable data is still not met. To further improve diabetic complication prediction models, biomarkers were noted to be especially beneficial [5].

In this study we evaluate an established biomarker of aging, DNA methylation age acceleration (DNAmAA) [21], in a sample of 1,100 participants of the Berlin Aging Study II (BASE-II) that were reassessed on average 7.4 years later as part of the GendAge study. This large longitudinal cohort has been investigated with respect to prevalence and incidence of type 2 diabetes (T2D) before ([22]).

In a first step, the cross-sectional relationship between DNAmAA estimated from five epigenetic clocks (7-CpG clock, Horvath’s clock, Hannum’s clock, PhenoAge, GrimAge), prevalent T2D, and several diabetes-associated blood parameters was examined. Previously published studies reported contradictory results. Dugue and colleagues [23], Irvin and colleagues [24], and Roetker and colleagues [25] found statistically significant associations between Horvath and Hannum clock estimates and T2D. However, McCartney and colleagues [26] and Horvath and colleagues [27] did not find any association between both variables. In a second step, we analyzed these epigenetic clocks with respect to diabetes associated complications which were measured with the Diabetes Complications Severity Index (DCSI). To our knowledge, this is the first study that explores this relationship.

Cross-sectional associations were found for the Hannum and PhenoAge DNAmAA and longitudinal analyses suggest that the 7-CpG DNAmAA can help in the prediction of diabetic complications in men.

## Methods

### BASE-II and GendAge study

The BASE-II is an explorative multi-disciplinary study that examines factors promoting “healthy” vs. “unhealthy” aging. The medical part of this study included 1,671 participants of the greater Berlin metropolitan area between the age of 60-85 years as assessed at baseline between 2009 and 2014. An additional group of 500 younger participants was assessed as well (age range 20 to 37 years) but is not analyzed in this study. After an average follow-up period of 7.4 years, 1,083 participants of the older age group were reassessed as part of the GendAge study and are therefore available for longitudinal analyses. An additional 17 participants were only assessed at follow-up examination.

All participants gave written informed consent. The medical assessments at baseline and follow-up were conducted in accordance with the Declaration of Helsinki and approved by the Ethics Committee of the Charité – UniversitaCtsmedizin Berlin (approval numbers EA2/029/09 and EA2/144/16) and were registered in the German Clinical Trials Registry as DRKS00009277 and DRKS00016157.

### DNA methylation age

DNA methylation age (DNAmA) estimated from the 7-CpG clock was available for baseline and follow-up examination. This clock was trained on samples obtained from BASE-II participants during baseline examination [28] and is calculated from methylation data determined by methylation-sensitive single-nucleotide primer extension (MS-SNuPE) [29]. Briefly, genomic DNA was isolated from whole blood samples and bisulfite converted. Subsequently, the areas of interest were amplified by a multiplex polymerase chain reaction. Finally, the SNuPE reaction was performed and the methylation fraction of the seven CpG sites of interest was measured by a “3730 DNA Analyzer” (Applied Biosystems, HITACHI). The average change of 7-CpG DNAmA between baseline and follow up per year was 0.75 years (SD = 0.64 years, range: -4.8 to 5.4 years, n=965, [30]) and correlation between DNAmA at T0 and T1 was high (r= Pearson’s r=0.81, n=965, [30]). A detailed description of methods was previously published for baseline [28] and follow-up [30, 31].

For follow-up only, four additional DNAmA measures were available that were estimated from epigenome wide DNAm profiles obtained through Illumina’s “Infinium MethylationEPIC” array (Illumina Inc., USA). Horvath’s clock [32], Hannum’s clock [33], PhenoAge [34] and GrimAge [35] were used to estimate DNAmA by the manual on Steve Horvath’s website (https://horvath.genetics.ucla.edu/html/dnamage/). Briefly, outliers (identified by the outlyx and pcout function in the R-package “bigmelon” [36]) and samples with a bisulfite conversion efficiency below 80% were removed. The resulting sample set was reloaded and normalized with the package’s dasen function. Samples that had a root-mean squared deviation of 0.1 or more in beta-values before and after normalization were excluded. For epigenetic clock estimations, the raw (i.e., not normalized) DNAm values were uploaded to the website, according to the instructions in the manual. DNAmA of all five epigenetic clocks were moderately correlated with each other (Pearson’s r = 0.4 to 0.6, Supplementary Figure 2A of ref.[30]). Detailed information on methods and longitudinal and cross-sectional descriptive statistics of the available DNAmA measures at follow-up can be found in ref. [30].

**Figure 1:**
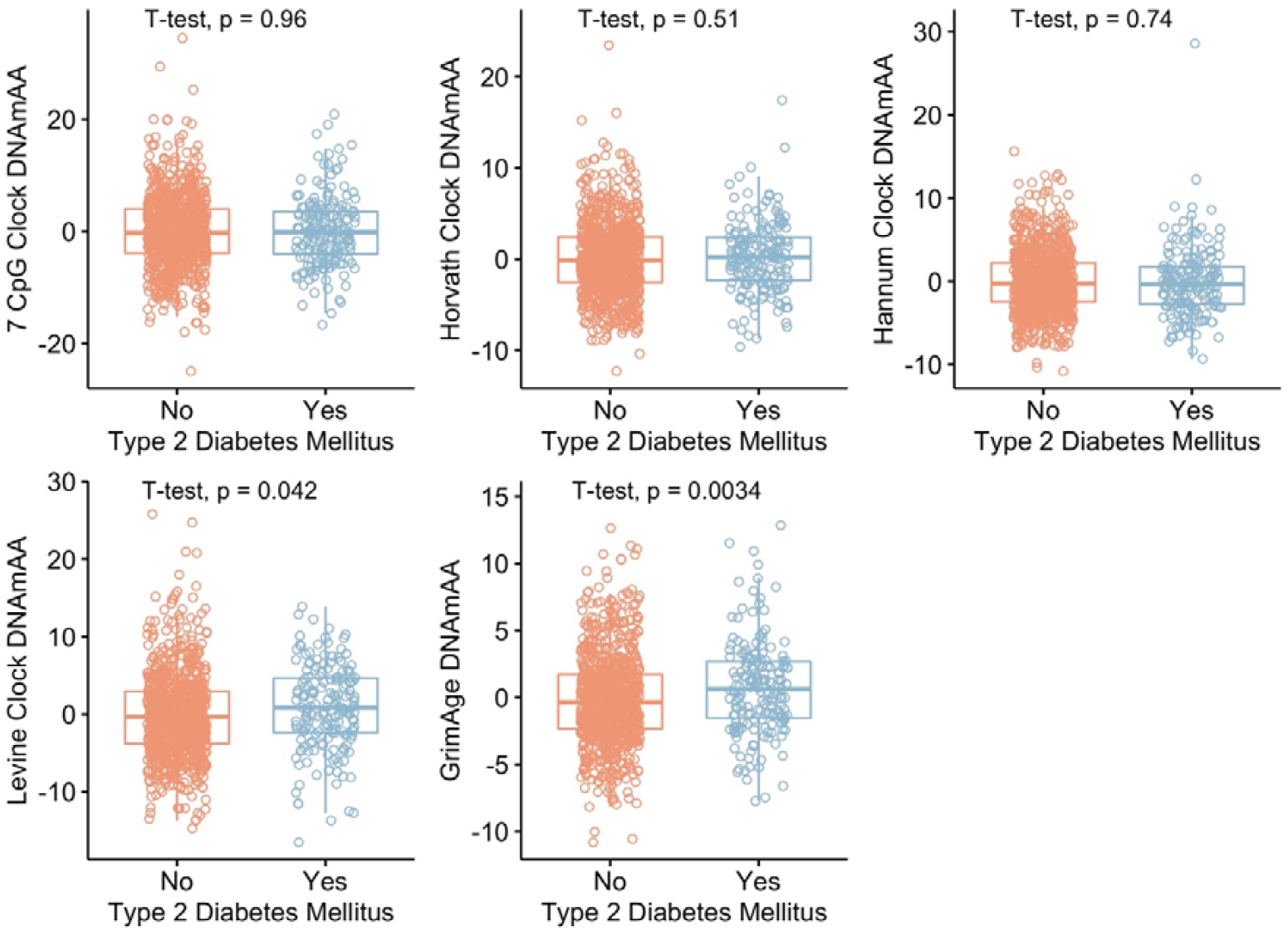
Boxplots of DNAmAA estimates by five different epigenetics clocks, stratified by their diagnostic status (T2D vs. no T2D).

**Figure 2:**
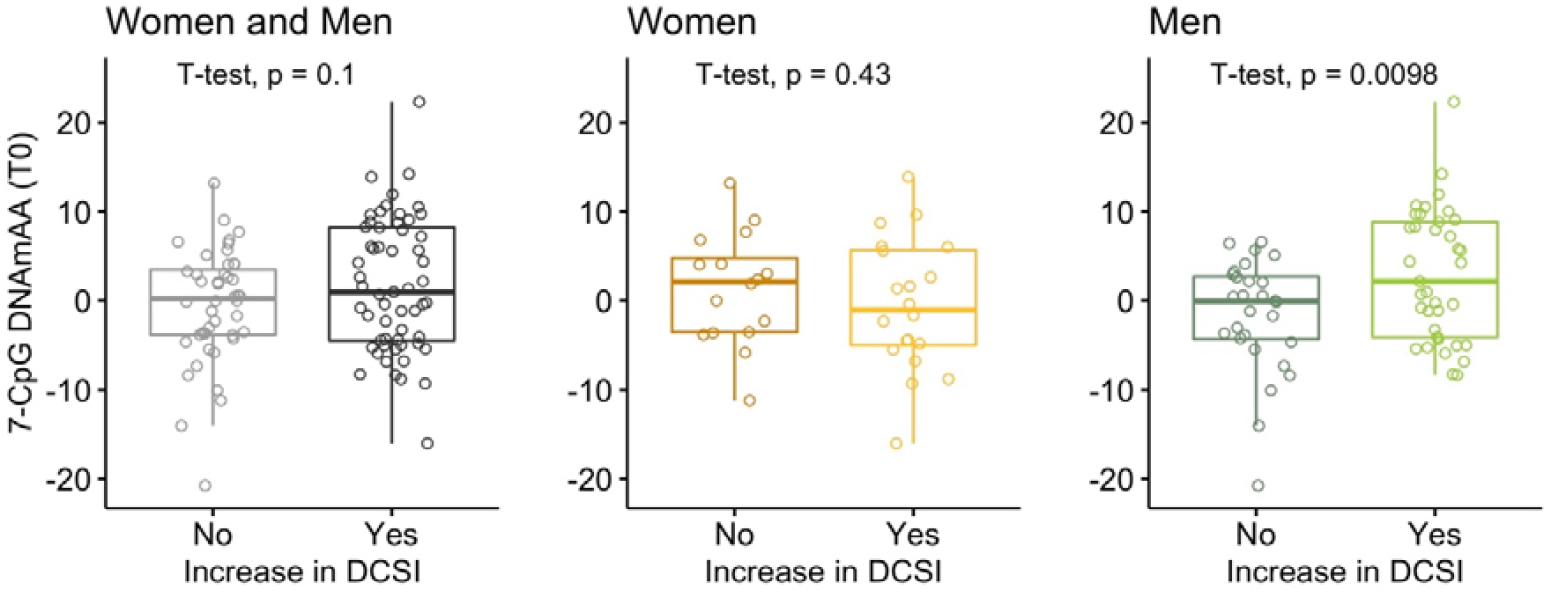
Boxplot of 7-CpG DNAmAA at baseline stratified by increase in DCSI during follow-up period. Analyses were performed on all participants and in sex-stratified subgroups.

### DNA methylation age acceleration

To adjust for known age-associated changes in leukocyte cell composition, we calculated DNAmAA as residuals from a cell-count (neutrophils, monocytes, lymphocytes, eosinophils) adjusted linear regression analysis of DNAmA on chronological age [28, 37]. All cell counts were determined in a certified routine laboratory by flow cytometry (MVZ Labor 28 GmbH, Berlin, Germany). As expected, correlation between DNAmAA and chronological age was low (|Pearson’s r|<0.09, [30]).

### Diabetes mellitus type 2 (T2D) and Diabetes Complications Severity Index (DCSI)

A detailed report on methodology and descriptive statistics of diabetes in BASE-II and GendAge was reported before ([22]). Diagnostic criteria of the American Diabetes Association (ADA) guidelines were used to identify participants with diabetes mellitus [38]. The DCSI which was developed by Young and colleagues [39] incorporates information about seven categories of complications: Retinopathy, nephropathy, neuropathy, cerebrovascular disease, cardiovascular disease, peripheral vascular disease, and metabolic complications. The resulting score is calculated based on the severity of the respective complication (0 = none, 1 = some and 2 = severe), except for neuropathy which can only result in a score of 0 or 1. Relevant information on the metabolic status were not available in this sample, therefore a maximum of 11 points was achievable. Dichotomized DCSI was calculated to distinguish the group of participants whose DCSI score did not increase (“not increased”) during follow-up from those whose DCSI score increased by at least one point (“increased”). The average change in DCSI per year during follow-up period was calculated as difference between DCSI at T1 and T0 divided by follow-up time. A detailed description on how the DCSI was assessed in this cohort is provided in ref. ([22]).

### Covariates

Sex-differences in aging and in DNAmAA are well documented. Therefore, sex was included in all regression models and sex-stratified analyses were performed. Smoking behavior (in packyears) and alcohol consumption (baseline: g/d via food frequency questionnaire [40], follow-up: yes/no) were assessed in one-to-one interviews. Body mass index was calculated by height and weight measures obtained from the “763 seca” measuring station (SECA, Germany). Diabetic medication was assessed by participants self-reports and/or from the medication list participants provided during both examinations.

### Statistical Analyses

Statistical analyses and all figures were conducted in R 3.6.2 [41]. ROC curves were calculated with the *proc* package [42]. Significance of difference between ROC curves was computed with the “*roc.test”* [42] function that employs the approach described by DeLong and colleagues [43].

An available case analysis was performed. Therefore, participants who did not provide information on all variables needed for an analysis were excluded from it. The analyzed sample size is indicated for each analysis. Nominal statistical significance was defined at an alpha of 0.05.

## Results

### Cohort characteristics

As previously reported, 12.9% (n=209 of a total sample of n=1,625) of participants were diagnosed with T2D at baseline ([22]). On average, 7.4 years later this number increased to 17.1% (n=185 of a total sample of n=1,083) among the participants who completed the follow-up assessments [22].

The mean age of BASE-II participants with diagnosed T2D at baseline and a completed follow-up examination was 68.0 years (SD= 3.7 years, n=126, 41.3 % female, Table 1) at baseline and 75.5 years (SD= 4.1 years, Table 1) at follow-up. The DCSI of 54.8% (n=69) of these participants increased during the on average 7.4-year follow-up period. The average increase in this group was 2.5 points for women (SD = 1.4, n= 28) and 1.9 points in the subgroup of men (SD = 1.0, n=41, data not shown). Male participants with T2D were found to have a significantly higher DCSI than women with T2D at baseline (difference = 0.71 points, p=0.005, Supplementary Table 1). This difference was smaller and not statistically significant at follow-up (difference = 0.26, p=0.5, Supplementary Table 1). Approximately the same number of men and women had a higher DCSI score at follow-up compared to baseline (women: 53.9%, men: 55.4%, Supplementary Table 1).

**Table 1:** Cohort characteristic of participants of the older age group that were diagnosed with type 2 diabetes at baseline and completed follow-up on average 7.4 years later (n=126).

|  | n | mean, % | sd | min | max |
| --- | --- | --- | --- | --- | --- |
| Sex (female) | 52 | 41.27 |  |  |  |
| Chronological Age (T0) | 126 | 68.04 | 3.68 | 61.37 | 77.29 |
| Chronological Age (T1) | 126 | 75.46 | 4.09 | 66.78 | 85.94 |
| 7-CpG DNAmA (T0) | 109 | 66.68 | 8.00 | 40.25 | 90.62 |
| 7-CpG DNAmAA (T0) | 101 | 0.75 | 7.09 | -20.75 | 22.33 |
| Smoking (packyears, T0) | 101 | 15.41 | 18.77 | 0 | 80 |
| Alcohol intake (g/d, T0) | 108 | 15.75 | 16.65 | 0.44 | 86.40 |
| BMI (T0) | 110 | 29.40 | 4.15 | 19.41 | 39.87 |
| Antidiabetic medication<br>(yes, T0) | 66 | 52.80 |  |  |  |
| DCSI (T0) | 126 | 1.13 | 1.40 | 0 | 7 |
| DCSI (T1) | 126 | 2.07 | 1.87 | 0 | 7 |
| DCSI increase (yes) | 69 | 54.76 |  |  |  |
| Fasting Glucose (T0) | 121 | 123.45 | 31.61 | 59.00 | 254.00 |
| oGTT (T0)* | 42 | 193.64 | 61.45 | 94.00 | 308.00 |
| HbA1c (T0) | 119 | 6.48 | 0.73 | 5.00 | 8.90 |

Note: DNAmA = DNA methylation age, DNAmAA = DNA methylation age acceleration; T0 = baseline examination, T1 = follow-up examination, g/d = gram per day, DCSI = Diabetes Complications Severity Index; oGTT = oral glucose tolerance test; HbA1c = glycated hemoglobin. \*OGTT was only assessed in participants that were not already diagnosed with diabetes before examination. Therefore, only comparatively few participants provide information in the subgroup shown here. Cohort characteristics of the whole dataset of participants at follow-up (n=1,100) are provided in Supplementary Table 5.

### Association between five DNAmAA measures, diagnosed diabetes mellitus and diagnostic blood parameters

We found participants with diagnosed diabetes mellitus to have 0.93 years higher PhenoAge DNAmAA (p=0.042, Figure 1) and 0.85 year higher GrimAge DNAmAA (p=0.003, Figure 1) than participant that were not diagnosed with diabetes mellitus. After adjustment for covariates in cross-sectional logistic regression analyses of the T2D diagnosis (yes/no) on DNAmAA of all five epigenetic clocks, no statistically significant association between these variables was found (*logistic regression*, Table 2, unadjusted models in Supplementary Table 2).

**Table 2:**
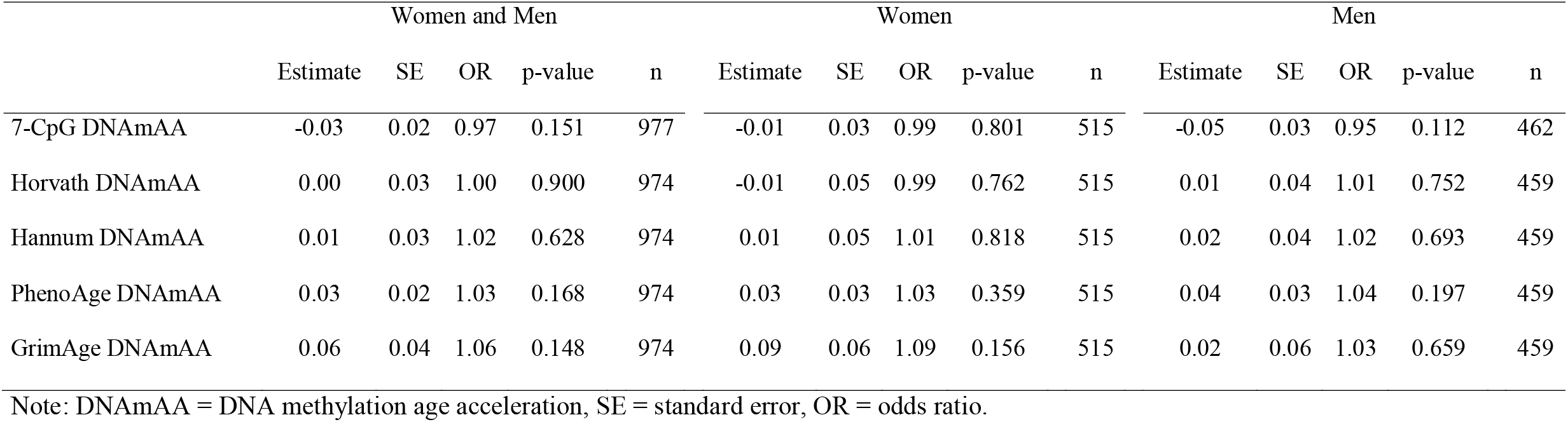
Logistic regression analyses of diagnosis of diabetes mellitus on DNAmAA of five different epigenetic clocks in cross-sectional data at follow-up. Analyses are adjusted for chronological age, sex (if applicable), alcohol consumption (yes/no), smoking (packyears), antidiabetic medication (yes/no), and BMI. All variables were assessed at follow-up. All available participants are included in these analyses.

The only available epigenetic clock estimate at baseline examination, 7-CpG DNAmAA, was neither cross-sectionally associated with diagnosed diabetes at baseline nor longitudinally associated with diagnosed diabetes at follow-up. This was true for sex-stratified analyses as well (*linear regression*, data not shown).

Fasting glucose was significantly associated with PhenoAge DNAmAA after adjustment for chronological age, sex, alcohol consumption (yes/no), smoking (packyears), diabetes medication (yes/no) and BMI (Table 3, β=0.26, SE=0.10, p=0.013, unadjusted models in Supplementary Table 3). Additionally, a statistically significant positive association was found between PhenoAge DNAmAA and results from the oral glucose tolerance test (oGTT, β=0.75, se=0.26, p=0.003, n=762, *linear regression*, Table 3). Similarly, a statistically significant association of about the same effect size was found between Hannum’s DNAmAA and oGTT in the highest adjusted model (β=0.78, se=0.34, p=0.023, n=762, *linear regression*, Table 2). However, none of the examined DNAmAA measures were associated with glycosylated hemoglobin (HbA1c, p>0.2, Table 3).

**Table 3:** Linear regression analyses of continuous blood parameters on DNAmAA of five epigenetic clocks in cross-sectional data at follow-up. Analyses are adjusted for chronological age, sex, alcohol consumption (yes/no), smoking (packyears), antidiabetic medication (yes/no), and BMI. All variables were assessed at follow-up. All available participants were included in this analysis.

|  | Fasting Glucose |  |  |  | 2h-oGTT |  |  |  | HbA1c |  |  |  |
| --- | --- | --- | --- | --- | --- | --- | --- | --- | --- | --- | --- | --- |
|  | Estimate | SE | p-value | n | Estimate | SE | p-value | n | Estimate | SE | p-value | n |
| 7-CpG DNAmAA | 0.04 | 0.09 | 0.677 | 969 | 0.11 | 0.21 | 0.596 | 765 | 0.00 | 0.00 | 0.934 | 973 |
| Horvath DNAmAA | 0.19 | 0.14 | 0.161 | 966 | 0.35 | 0.33 | 0.300 | 762 | 0.00 | 0.00 | 0.915 | 970 |
| Hannum DNAmAA | 0.20 | 0.14 | 0.166 | 966 | 0.78 | 0.34 | 0.023 * | 762 | 0.00 | 0.00 | 0.314 | 970 |
| PhenoAge DNAmAA | 0.26 | 0.10 | 0.013 * | 966 | 0.75 | 0.26 | 0.003 ** | 762 | 0.00 | 0.00 | 0.201 | 970 |
| GrimAge DNAmAA | 0.25 | 0.19 | 0.183 | 966 | 0.01 | 0.46 | 0.980 | 762 | 0.00 | 0.00 | 0.441 | 970 |

Note: DNAmAA = DNA methylation age acceleration, SE = standard error, HbA1c = glycated hemoglobin, oGTT = oral glucose tolerance test.

### Cross-sectional association between DNAmAA of five different epigenetic clocks and DCSI

Cross-sectional analyses between 7-CpG DNAmAA and DCSI at baseline examination did not reveal statistically significant associations (Supplementary Table 4). Additional to the longitudinally available 7-CpG clock, four more epigenetic clocks were available at follow-up examination. Cohort characteristics of analyzed participants at follow-up are displayed in Supplementary Table 5. Cross-sectional linear regression analyses were performed to analyze the relationship between these biological age measures and DCSI in participants with diagnosed T2D at follow-up. No association between any of the available epigenetic clocks and DCSI was statistically significant after adjustment for covariates (*linear regression*, Table 4). Unadjusted and minimally adjusted models are displayed in Supplementary Table 6.

**Table 4:** Linear regression analyses of DCSI on DNAmAA and covariates in cross-sectional data at follow-up. Included are participants with diagnosed diabetes mellitus at follow-up. Analyses are adjusted for chronological age, sex (if applicable), alcohol consumption (yes/no), smoking (packyears), antidiabetic medication (yes/no), and BMI. All variables were assessed at follow-up.

|  | Women and Men |  |  |  | Women |  |  |  | Men |  |  |  |
| --- | --- | --- | --- | --- | --- | --- | --- | --- | --- | --- | --- | --- |
|  | Estimate | SE | p-value | n | Estimate | SE | p-value | n | Estimate | SE | p-value | n |
| 7-CpG DNAmAA | 0.04 | 0.02 | 0.087 | 163 | 0.03 | 0.03 | 0.372 | 67 | 0.05 | 0.03 | 0.114 | 96 |
| Horvath DNAmAA | -0.03 | 0.04 | 0.377 | 163 | -0.04 | 0.06 | 0.488 | 67 | -0.01 | 0.04 | 0.787 | 96 |
| Hannum DNAmAA | 0.00 | 0.03 | 0.974 | 163 | 0.07 | 0.06 | 0.264 | 67 | -0.03 | 0.04 | 0.479 | 96 |
| PhenoAge DNAmAA | 0.03 | 0.03 | 0.302 | 163 | 0.00 | 0.04 | 0.945 | 67 | 0.02 | 0.03 | 0.474 | 96 |
| GrimAge DNAmAA | -0.06 | 0.05 | 0.208 | 163 | -0.14 | 0.09 | 0.152 | 67 | -0.04 | 0.06 | 0.473 | 96 |

Note: DNAmAA = DNA methylation age acceleration, SE = standard error.

**Table 5:** Logistic regression of increase in DCSI of one or more points vs. no increase in DCSI (dichotomized) between baseline and follow-up on 7-CpG DNAmAA at baseline and covariates. Analyses are adjusted for DCSI (T0), chronological age (T0), sex (if applicable), smoking (packyears, T0), alcohol consumption (g/d, T0), antidiabetic medication (yes/no, T0), and BMI (T0). Included are all participants with diagnosed diabetes mellitus at baseline.

| Model | Estimate | SE | OR | p-value | n |
| --- | --- | --- | --- | --- | --- |
| All | 0.040 | 0.033 | 1.041 | 0.219 | 89 |
| Women | -0.004 | 0.061 | 0.996 | 0.949 | 33 |
| Men | 0.101 | 0.051 | 1.106 | 0.045 | * 56 |

Note: T0 = baseline, SE = standard error, OR = odds ratio.

### 7-CpG DNAmAA predicts additional complications after 7.4-year follow-up time in subgroup of men

Participants who reported additional or worsened complications during the follow-up period had a 2.3 year higher 7-CpG DNAmAA at baseline (p=0.1, Figure 2). Sex-stratified subgroup analyses revealed a statistically significant difference of 4.5 years higher baseline 7-CpG DNAmAA in men whose DCSI increased after examination (p<0.001, Figure 2). However, in the female subgroup this association was inverse and not statistically significant (difference between means: 1.8 years, p=0.4, Figure 2).

To account for potential confounders, we performed a logistic regression analysis of the dichotomized change in DCSI on 7-CpG DNAmAA and covariates. In line with the findings reported above, we found a 11% increase of the risk for developing at least one additional or worsened complication captured by the DCSI during follow-up period for every additional year of DNAmAA in the subgroup of men. This association was independent from DCSI at baseline, chronological age, smoking, alcohol consumption, diabetes medication and BMI (OR = 1.11, p=0.045, n=56, *logistic regression*, Table 5, unadjusted models in Supplementary Table 7). No statistically significant association was found in the whole group (OR=1.04, p=0.22) and in the female subgroup (OR=0.996, p=0.95, Table 5). Similar results were found when analyzing the continuous change in DCSI per year during the follow-up period in a linear regression analysis (Supplementary Table 8). Receiver operating characteristic (ROC) curves of the highest adjusted model are displayed in Figure 3. The rea under the curve (AUC) of the ROC curve that resulted from the logistic regression model of all participants and the male and female subgroups was 0.6, 0.8 and 0.7. Interestingly, the inclusion of DNAmAA improved the predictive model in the subgroup of women of about the same degree as in the male subgroup (Figure 3), although it did not contribute significantly to the logistic regression model. This might be the result of the small sample size available in this subgroup. The ROC-curves did not differ statistically (p>0.05) which probably needs to be attributed to the small sample size as well.

**Figure 3:**
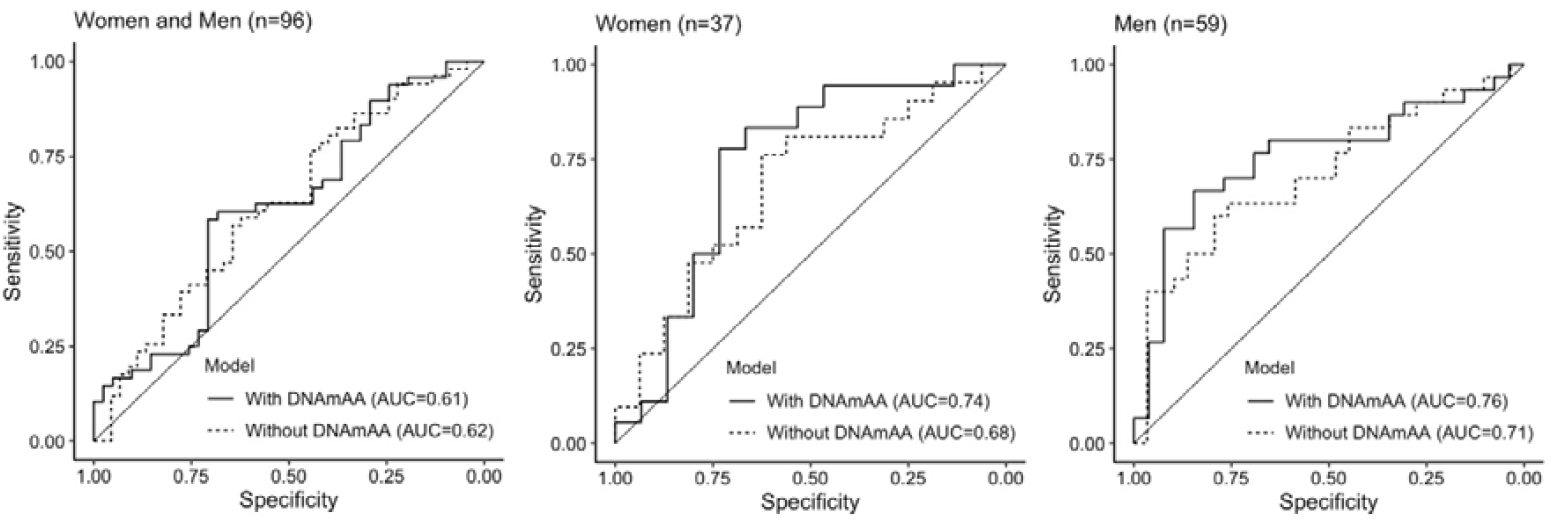
Prediction of T2D complications as assessed with the DCSI. The basic model used for complication prediction includes chronological age (years), sex (if applicable), alcohol intake (g/d), packyears (years), diabetes medication (yes/no), and BMI. The differences between ROC curves of the model with and without 7-CpG DNAmAA were statistically not significant (p>0.05).

## Discussion

As a main result of the current study, we found that the 7-CpG clock derived variable DNAmAA was not associated with the diagnosis of T2D. However, baseline 7-CpG DNAmAA was associated with T2D severity on average 7.4years later as operationalized by the DCSI in men. Although the four other analyzed epigenetic clocks were only available for cross-sectional analyses at follow-up, a similar predictive ability would be expected at least for PhenoAge DNAmAA. This clock derivate was associated with diagnosis of T2D, fasting glucose and results in the oGTT, a well-established test for impaired glucose tolerance. These promising findings suggest that the 7-CpG clock (and possibly other clocks as well) might be able to substantially improve future diabetic complication risk scores.

The predictive ability of 7-CpG DNAmAA was analyzed by AUC of the logistic regression models of increase in DCSI (yes/no) during follow-up on 7-CpG DNAmAA at baseline and covariates which was in the acceptable range (AUC = 0.8, Figure 3) [44]. To our knowledge, there are no models available that aim at the prediction of the DCSI and could be used for a meaningful comparison of performance. However, numerous scores for specific micro- and macrovascular complications were reported before (reviewed in ref. [12-14, 16, 18]). Saputro and colleagues reported on pooled C-statistics of logit-based models from cohort studies. Results for diabetic nephropathy (11 studies, 1 to 10 years of follow-up time, AUC=0.78, [18]) and diabetic retinopathy (6 studies, 1 to 20 years of follow-up time, AUC=0.82, [18]) were in a similar range to the model reported in this study. Similarly, Chowdhury and colleagues reported on a median AUC of 0.71 of nine models designed to predict stroke in patients with diabetes mellitus (1.4 to 10.5 years of follow-up time, [14]) and Beulens and colleagues reported on an AUC between 0.54 and 0.81 of 20 models that aimed at the prediction of foot ulcer (1 to 5 years follow-up time, [45]). A detailed analysis of the previously developed risk models and the incorporated predictors is beyond the scope of this manuscript and was done before [12-14, 16, 18] and the comparability between models is limited due to the high variability in the methods, cohorts and outcomes that were used to develop these risk scores. However, the results show that the 7-CpG DNAmAA informed risk model presented in this study performs in a comparable range (and sometimes better) than more complication specific risk models. This is especially intriguing as several aspects of this still comparatively new biomarker seem very well suited for prediction modelling. First, the 7-CpG clock can be obtained through two different methodological approaches. It was developed for the MS-SNuPE method that can be conducted cost-effectively even in smaller laboratories [28], but additionally can be determined based on epigenome-wide array data, such as data obtained by microarrays [31]. Therefore, it can easily be applied to cohorts where epigenome-wide data is already available. Second, biomarkers allow an objective assessment of the individual complication risk. They are independent from factors that might interfere with data assessment in a clinical context, such as a language barrier, inaccurate or biased memory, unstandardized documentation, or examiner bias. An advantage of this specific biomarker is that changes in the epigenetic clock seem to result from lifelong and cumulative influences [46]. Therefore, it is expected to be robust against short-term changes and circadian differences that can complicate the use of biomarkers in a clinical context. In contrast to (poly-)genetic risk scores that are set at birth, the epigenetic clock changes throughout life. We and others have shown that these changes are potentially sensitive to interventions [47-51] and lifestyle factors [37]. Whether this is true for diabetes specific interventions needs to be examined in sufficiently sized longitudinal studies. A meaningful relationship, however, seems plausible because high levels of glucose were shown to change DNA methylation by upregulating methylating enzymes and downregulating demethylating enzymes in in-vitro experiments of rat cells [52].

Despite these promising results in terms of complications related to diabetes, no association was found between diabetes diagnosis and DNAmAA of all five available clocks after adjustment for covariates (Table 2). In contrast to the first-generation clocks (7-CpG, Horvath’s and Hannum’s clock) that were trained to predict chronological age, PhenoAge and GrimAge were trained to predict phenotype-based biological age estimates. The negative findings with respect to the first-generation clocks reported here are in line with results that were reported by [26] McCartney and colleagues and Horvath and colleagues [27]. In contrast, positive associations between Horvath’s or Hannum’s DNAmAA and diabetes were reported by Dugue and colleagues [23], Irvin and colleagues [24], and Roetker and colleagues [25]. However, these cohorts differed in sex-distribution, age-range, or their statistical approach from the analyses presented here, which might at least partially explain the difference in findings. To our knowledge, GrimAge is the only second-generation clock that was examined with respect to T2D before. Kim and colleagues found an association between GrimAge DNAmAA and T2D (OR=2.57, 95% CI: 1.61-4.11) in 318 obese participants (mean age = 40 years, 53% female). This association could not be found in the overweight and normal weight group of the study. An association between GrimAge DNAmAA and T2DM was found in this study as well (OR=1.07, p=0.002, n=1051, *model 1*, Supplementary Table 2), but in contrast to the findings by Kim and colleagues this association did not persist after inclusion of covariates.

To further evaluate the epigenetic clocks in the context of diabetes, we performed linear regression analyses of diabetes associated blood parameters on DNAmAA. A statistically significant association between PhenoAge DNAmAA and fasting glucose was found (Table 3). This association seems plausible because serum glucose was included in the phenotypic age measure that was used to train the PhenoAge clock [34]. The oGTT, a test used to assess how glucose is metabolized and that is used to diagnose T2D, was significantly associated with PhenoAge DNAmAA and Hannum’s DNAmAA after covariate adjustment (Table 3). To our knowledge, this association has not been examined before.

There are several limitations to this study which we summarize as follows. First, the small sample size of participants with diagnosed diabetes mellitus at baseline might be the reason for the lack of statistical significance in some analyses. A replication of these analyses in larger cohorts of patients with diabetes is therefore needed. Second, we were not able to evaluate the predictive ability of the logistic regression model in an independent dataset. However, such independent validation and calibration analyses are crucial before translation into clinical practice is possible. Third, due to the exploratory approach of this study and in line with most studies of this field, we did not adjust our analyses for multiple testing. However, an increased rate of false-positive findings can therefore not be ruled out and our findings need to be replicated and validated in an independent cohort. Fourth, the outcome variable that represents diabetic complications, DCSI, reflects a general burden instead of a highly differentiated assessments of a specific diabetes associated complication. However, the general assessment of risk for complications is closer to clinical practice than a potentially better but complication-specific risk estimation. This could help implementing a risk score in clinical practice in the future, where health care workers are challenged with the identification of patients with a generally high risk for complications rather than a high risk for specific complications.

Strengths of this study include the wide variety of different epigenetic clock estimates assessed and the longitudinally available 7-CpG clock, which allowed a comprehensive evaluation of this still comparatively new biomarker of aging. Furthermore, the analyzed BASE-II cohort is well characterized with respect to diabetes mellitus [22] and provides a robust data base that allows the comparison of different diabetes-associated variables and adjustment for relevant covariates.

## Conclusion

None of the epigenetic clocks analyzed in this study were associated with diagnosis of T2D after adjustment for covariates. However, Hannum and PhenoAge DNAmAA showed a statistically significant association with oGTT. Furthermore, PhenoAge DNAmAA was associated with fasting glucose. Despite the lack of association between the 7-CpG clock with T2D and associated blood parameters, a one-year increase in 7-CpG DNAmAA was associated with 11% increase in the risk for development of additional T2D-related complications on average 7.4 years later in men. Although this association still lacks external validation, the results suggest that DNAmAA as a biomarker might be able to improve the identification of the group of diabetes patients that is especially prone to complications. Therefore, already existing prediction models might be substantially improved by the inclusion of 7-CpG DNAmAA which ultimately might accelerate their translation to clinical practice.

## Data Availability

Data are available upon reasonable request. Interested investigators are invited to contact the study coordinating PI Ilja Demuth at to obtain additional information about the GendAge study and the data-sharing application form.

## Acknowledgments

-

## Author contributions

Conceived and designed the study: VMV and ID. Contributed study specific data: all authors. Analyzed the data: VMV and YS. Wrote the manuscript: VMV and ID. All authors revised and approved the manuscript.

