## Supplementary Material for "DNA Methylation Age Acceleration, Type 2 Diabetes, and its Complications: Cross-sectional and Longitudinal Data from the Berlin Aging Study II (BASE-II)"

**Corresponding author:**

**Supplementary Table 1: Sex-stratified cohort characteristics of participants with diagnosed diabetes at baseline examination (n=126).** Significance of differences between sexes was assessed by t-test and chi-squared test.

| | Women | | | | | Men | | | | | |
|---|---|---|---|---|---|---|---|---|---|---|---|
| | n | Mean, % | SD | Min | Max | n | Mean, % | SD | Min | Max | p-value |
| Chronological Age (T0) | 52 | 68.09 | 3.56 | 61.91 | 76.95 | 74 | 68.01 | 3.78 | 61.37 | 77.29 | 0.904 |
| Chronological Age (T1) | 74 | 75.26 | 4.27 | 66.78 | 85.94 | 52 | 75.74 | 3.83 | 68.91 | 85.61 | 0.520 |
| 7-CpG DNAmA (T0) | 41 | 66.08 | 8.24 | 49.31 | 89.03 | 68 | 67.05 | 7.89 | 40.25 | 90.62 | 0.540 |
| 7-CpG DNAmAA (T0) | 36 | 0.37 | 6.89 | -16.01 | 13.90 | 65 | 0.95 | 7.24 | -20.75 | 22.33 | 0.694 |
| Smoking (packyears, T0) | 39 | 7.72 | 13.47 | 0 | 45 | 62 | 20.24 | 20.08 | 0 | 80 | 0.001 |
| Alcohol intake (g/d, T0) | 40 | 12.10 | 14.98 | 0.62 | 65.16 | 68 | 17.90 | 17.30 | 0.44 | 86.40 | 0.081 |
| BMI (T0) | 43 | 29.60 | 4.54 | 21.04 | 39.87 | 67 | 29.27 | 3.91 | 19.41 | 39.62 | 0.684 |
| Diabetes medication (yes, T0) | 25 | 49.02 | | | | 41 | 55.41 | | | | 0.603 |
| DCSI (T0) | 52 | 0.71 | 1.26 | 0 | 7 | 74 | 1.42 | 1.42 | 0 | 5 | 0.005 |
| DCSI (T1) | 52 | 1.92 | 1.90 | 0 | 7 | 74 | 2.18 | 1.85 | 0 | 6 | 0.457 |
| DCS increase (yes) | 28 | 53.85 | | | | 41 | 55.41 | | | | 1.000 |
| Fasting Glucose (T0) | 49 | 114.45 | 23.70 | 69.00 | 158.00 | 72 | 129.57 | 34.86 | 59.00 | 254.00 | 0.009 |
| oGTT (T0) | 20 | 175.75 | 62.21 | 94.00 | 308.00 | 22 | 209.91 | 57.36 | 94.00 | 307.00 | 0.071 |
| HbA1c (T0) | 48 | 6.40 | 0.70 | 5.00 | 8.70 | 71 | 6.54 | 0.74 | 5.20 | 8.90 | 0.299 |

Note: DNAmA = DNA methylation age, DNAmAA = DNA methylation age acceleration; T0 = baseline examination, T1 = follow-up examination, g/d = gram per day, BMI = body mass index, DCSI = Diabetes Complications Severity Index; oGTT = oral glucose tolerance test; HbA1c = glycated hemoglobin.

**Supplementary Table 2: Logistic regression analyses of diagnosis of diabetes mellitus on DNAmAA of five different epigenetic clocks in cross-sectional data at follow-up.** Model 1: no covariates; Model 2: chronological age, sex (if applicable), alcohol consumption (yes/no), smoking (packyears), antidiabetic medication (yes/no), BMI. All variables were assessed at follow-up. All available participants are included in these analyses.

| | | Women and Men | | | | | | Women | | | | | Men | | | | |
|---|---|---|---|---|---|---|---|---|---|---|---|---|---|---|---|---|---|
| | Model | Estimate | SE | OR | p-value | | n | Estimate | SE | OR | p-value | n | Estimate | SE | OR | p-value | n |
| 7-CpG DNAmAA | 1 | 0.00 | 0.01 | 1.00 | 0.959 | | 1055 | 0.02 | 0.02 | 1.02 | 0.336 | 549 | -0.03 | 0.02 | 0.97 | 0.096 | 506 |
| | 2 | -0.03 | 0.02 | 0.97 | 0.151 | | 977 | -0.01 | 0.03 | 0.99 | 0.801 | 515 | -0.05 | 0.03 | 0.95 | 0.112 | 462 |
| Horvath DNAmAA | 1 | 0.01 | 0.02 | 1.01 | 0.516 | | 1051 | 0.02 | 0.03 | 1.02 | 0.583 | 549 | 0.00 | 0.03 | 1.00 | 0.926 | 502 |
| | 2 | 0.00 | 0.03 | 1.00 | 0.900 | | 974 | -0.01 | 0.05 | 0.99 | 0.762 | 515 | 0.01 | 0.04 | 1.01 | 0.752 | 459 |
| Hannum DNAmAA | 1 | -0.01 | 0.02 | 0.99 | 0.720 | | 1051 | 0.00 | 0.03 | 1.00 | 0.934 | 549 | -0.04 | 0.03 | 0.96 | 0.216 | 502 |
| | 2 | 0.01 | 0.03 | 1.02 | 0.628 | | 974 | 0.01 | 0.05 | 1.01 | 0.818 | 515 | 0.02 | 0.04 | 1.02 | 0.693 | 459 |
| PhenoAge DNAmAA | 1 | 0.03 | 0.01 | 1.03 | 0.038 | * | 1051 | 0.03 | 0.02 | 1.03 | 0.249 | 549 | 0.03 | 0.02 | 1.03 | 0.185 | 502 |
| | 2 | 0.03 | 0.02 | 1.03 | 0.168 | | 974 | 0.03 | 0.03 | 1.03 | 0.359 | 515 | 0.04 | 0.03 | 1.04 | 0.197 | 459 |
| GrimAge DNAmAA | 1 | 0.07 | 0.02 | 1.07 | 0.002 | ** | 1051 | 0.03 | 0.04 | 1.03 | 0.538 | 549 | 0.06 | 0.03 | 1.06 | 0.066 | 502 |
| | 2 | 0.06 | 0.04 | 1.06 | 0.148 | | 974 | 0.09 | 0.06 | 1.09 | 0.156 | 515 | 0.02 | 0.06 | 1.03 | 0.659 | 459 |

Note: DNAmAA = DNA methylation age acceleration, SE = standard error, OR = odds ratio.

**Supplementary Table 3: Linear regression analyses of blood parameters on DNAmAA of five epigenetic clocks in cross-sectional data at follow-up.** Model 1: no covariates; Model 2: chronological age, sex, alcohol consumption (yes/no), smoking (packyears), antidiabetic medication (yes/no), BMI. All variables were assessed at follow-up. All available participants were included in this analysis.

| | | Fasting Glucose | | | | | 2h-oGTT | | | | | HbA1c | | | |
|---|---|---|---|---|---|---|---|---|---|---|---|---|---|---|---|
| | Model | Estimate | SE | p-value | | n | Estimate | SE | p-value | | n | Estimate | SE | p-value | n |
| 7-CpG DNAmAA | 1 | 0.16 | 0.11 | 0.135 | | 1046 | -0.03 | 0.20 | 0.901 | | 823 | 0.00 | 0.00 | 0.498 | 1051 |
| | 2 | 0.04 | 0.09 | 0.677 | | 969 | 0.11 | 0.21 | 0.596 | | 765 | 0.00 | 0.00 | 0.934 | 973 |
| Horvath DNAmAA | 1 | 0.29 | 0.17 | 0.092 | | 1042 | 0.07 | 0.32 | 0.833 | | 820 | 0.00 | 0.00 | 0.724 | 1047 |
| | 2 | 0.19 | 0.14 | 0.161 | | 966 | 0.35 | 0.33 | 0.300 | | 762 | 0.00 | 0.00 | 0.915 | 970 |
| Hannum DNAmAA | 1 | 0.13 | 0.18 | 0.470 | | 1042 | 0.58 | 0.33 | 0.080 | | 820 | -0.01 | 0.00 | 0.085 | 1047 |
| | 2 | 0.20 | 0.14 | 0.166 | | 966 | 0.78 | 0.34 | 0.023 | * | 762 | 0.00 | 0.00 | 0.314 | 970 |
| PhenoAge DNAmAA | 1 | 0.44 | 0.13 | 0.001 | *** | 1042 | 0.68 | 0.24 | 0.005 | ** | 820 | 0.01 | 0.00 | 0.071 | 1047 |
| | 2 | 0.26 | 0.10 | 0.013 | * | 966 | 0.75 | 0.26 | 0.003 | ** | 762 | 0.00 | 0.00 | 0.201 | 970 |
| GrimAge DNAmAA | 1 | 0.81 | 0.20 | <0.001 | *** | 1042 | -0.06 | 0.38 | 0.877 | | 820 | 0.01 | 0.00 | 0.055 | 1047 |
| | 2 | 0.25 | 0.19 | 0.183 | | 966 | 0.01 | 0.46 | 0.980 | | 762 | 0.00 | 0.00 | 0.441 | 970 |

Note: DNAmAA = DNA methylation age acceleration, SE = standard error, HbA1c = glycated hemoglobin, oGTT = oral glucose tolerance test.

**Supplementary Table 4: Linear regression analyses of DCSI on DNAmAA and covariates in cross-sectional data at baseline.** Included are longitudinally available participants with diagnosed diabetes mellitus at baseline. Model 1: no covariates; Model 2: chronological age, sex (if applicable); Model 3: Model 2 + alcohol consumption (yes/no), smoking (packyears), antidiabetic medication (yes/no), + BMI. All variables were assessed at baseline.

| Model | Estimate | SE | p-value | n |
|---|---|---|---|---|
| | | Women and Men | | |
| 1 | 0.03 | 0.02 | 0.138 | 126 |
| 2 | 0.01 | 0.02 | 0.717 | 126 |
| 3 | 0.01 | 0.02 | 0.626 | 96 |
| | | Men | | |
| 1 | 0.03 | 0.02 | 0.175 | 74 |
| 2 | 0.01 | 0.02 | 0.678 | 74 |
| 3 | 0.02 | 0.03 | 0.399 | 59 |
| | | Women | | |
| 1 | 0.01 | 0.02 | 0.739 | 52 |
| 2 | 0.00 | 0.02 | 0.999 | 52 |
| 3 | -0.02 | 0.03 | 0.587 | 37 |

Note: SE = standard error.

**Supplementary Table 5: Descriptive statistics of all available participants at follow-up examination.** Significance of differences between sexes was assessed by t-test and chi-squared test.

| | Women and Men | | | | | Women | | | | | Men | | | | | |
|---|---|---|---|---|---|---|---|---|---|---|---|---|---|---|---|---|
| | n | Mean, % | SD | Min | Max | n | Mean, % | SD | Min | Max | n | Mean, % | SD | Min | Max | p-value |
| Chronological age (T1) | 1100 | 75.60 | 3.77 | 64.91 | 94.07 | 573 | 75.72 | 3.53 | 66.41 | 94.07 | 527 | 75.48 | 4.01 | 64.91 | 90.03 | 0.276 |
| Sex | | | | | | | | | | | | | | | | |
| male | 527 | 47.91 | | | | | | | | | 527 | 100.00 | | | | |
| female | 573 | 52.09 | | | | 573 | 100.00 | | | | | | | | | |
| Smoking (packyears, T1) | 1019 | 9.79 | 17.61 | 0.00 | 150.00 | 537 | 6.30 | 13.48 | 0.00 | 114.00 | 482 | 13.68 | 20.61 | 0.00 | 150.00 | <0.001 |
| Alcohol consumption (T1) | | | | | | | | | | | | | | | | |
| no | 185 | 16.86 | | | | 106 | 18.53 | | | | 79 | 15.05 | | | | 0.145 |
| yes | 912 | 83.14 | | | | 466 | 81.47 | | | | 446 | 84.95 | | | | |
| BMI (T1) | 1098 | 26.97 | 4.25 | 17.17 | 49.68 | 573 | 26.63 | 4.68 | 17.17 | 49.68 | 525 | 27.35 | 3.69 | 20.02 | 41.77 | 0.005 |
| 7-CpG DNAmAA (T1) | 1071 | 0.03 | 6.42 | -24.93 | 34.48 | 558 | -1.02 | 6.35 | -24.37 | 25.30 | 513 | 1.17 | 6.31 | -24.93 | 34.48 | <0.001 |
| Horvath DNAmAA (T1) | 1067 | 0.03 | 4.04 | -12.31 | 23.45 | 558 | -0.43 | 3.98 | -12.31 | 23.45 | 509 | 0.54 | 4.04 | -8.94 | 17.44 | <0.001 |
| Hannum DNAmAA (T1) | 1067 | 0.01 | 3.89 | -10.80 | 28.57 | 558 | -0.72 | 3.68 | -10.80 | 12.73 | 509 | 0.81 | 3.96 | -9.32 | 28.57 | <0.001 |
| PhenoAge DNAmAA (T1) | 1067 | 0.04 | 5.42 | -16.54 | 25.80 | 558 | -0.48 | 5.39 | -16.54 | 25.80 | 509 | 0.62 | 5.39 | -13.51 | 20.94 | 0.001 |
| GrimAge DNAmAA (T1) | 1067 | 0.03 | 3.39 | -10.82 | 12.84 | 558 | -1.30 | 2.93 | -10.82 | 10.71 | 509 | 1.47 | 3.27 | -8.17 | 12.84 | <0.001 |
| Fasting glucose (T1) | 1070 | 102.22 | 22.26 | 65.00 | 304.00 | 554 | 98.77 | 19.12 | 65.00 | 231.00 | 516 | 105.92 | 24.69 | 72.00 | 304.00 | <0.001 |
| oGTT (T1) | 837 | 118.46 | 37.39 | 32.00 | 358.00 | 442 | 117.94 | 35.59 | 44.00 | 244.00 | 395 | 119.04 | 39.34 | 32.00 | 358.00 | 0.671 |
| HbA1c (T1) | 1072 | 5.73 | 0.54 | 4.40 | 10.00 | 555 | 5.70 | 0.50 | 4.60 | 9.70 | 517 | 5.76 | 0.59 | 4.40 | 10.00 | 0.052 |
| DCSI (T1) | 1083 | 1.26 | 1.49 | 0.00 | 8.00 | 563 | 1.21 | 1.41 | 0.00 | 8.00 | 520 | 1.33 | 1.56 | 0.00 | 7.00 | 0.181 |
| Antidiabetic medication (T1) | | | | | | | | | | | | | | | | |
| no | 983 | 90.77 | | | | 528 | 93.78 | | | | 455 | 87.50 | | | | 0.001 |
| yes | 100 | 9.23 | | | | 35 | 6.22 | | | | 65 | 12.50 | | | | |
| Diagnosed diabetes mellitus (T1) | | | | | | | | | | | | | | | | |
| no | 896 | 82.89 | | | | 487 | 86.50 | | | | 409 | 78.96 | | | | 0.001 |
| yes | 185 | 17.11 | | | | 76 | 13.50 | | | | 109 | 21.04 | | | | |

Note: DNAmA = DNA methylation age, DNAmAA = DNA methylation age acceleration; T0 = baseline examination, T1 = follow-up examination, BMI = body mass index; DCSI = Diabetes Complications Severity Index; oGTT = oral glucose tolerance test; HbA1c = glycated hemoglobin.

**Supplementary Table 6: Linear regression analyses of DCSI on DNAmAA and covariates in cross-sectional data at follow-up.** Included are participants with diagnosed diabetes mellitus at follow-up. Model 1: no covariates; Model 2: chronological age, sex (if applicable); Model 3: Model 2 + alcohol consumption (yes/no), smoking (packyears), antidiabetic medication (yes/no), + BMI. All variables were assessed at follow-up.

| | | Women and Men | | | | | Women | | | | Men | | | | |
|---|---|---|---|---|---|---|---|---|---|---|---|---|---|---|---|
| | Model | Estimate | SE | p-value | | n | Estimate | SE | p-value | n | Estimate | SE | p-value | | n |
| 7-CpG DNAmAA | 1 | 0.05 | 0.02 | 0.029 | * | 180 | 0.03 | 0.03 | 0.387 | 73 | 0.06 | 0.03 | 0.040 | * | 107 |
| | 2 | 0.05 | 0.02 | 0.012 | * | 180 | 0.04 | 0.03 | 0.285 | 73 | 0.07 | 0.03 | 0.018 | * | 107 |
| | 3 | 0.04 | 0.02 | 0.087 | | 163 | 0.03 | 0.03 | 0.372 | 67 | 0.05 | 0.03 | 0.114 | | 96 |
| Horvath DNAmAA | 1 | -0.02 | 0.03 | 0.525 | | 179 | -0.05 | 0.06 | 0.408 | 73 | -0.01 | 0.04 | 0.800 | | 106 |
| | 2 | -0.01 | 0.03 | 0.680 | | 179 | -0.04 | 0.06 | 0.525 | 73 | 0.00 | 0.04 | 0.957 | | 106 |
| | 3 | -0.03 | 0.04 | 0.377 | | 163 | -0.04 | 0.06 | 0.488 | 67 | -0.01 | 0.04 | 0.787 | | 96 |
| Hannum DNAmAA | 1 | 0.02 | 0.03 | 0.491 | | 179 | 0.06 | 0.06 | 0.343 | 73 | 0.00 | 0.04 | 0.915 | | 106 |
| | 2 | 0.01 | 0.03 | 0.742 | | 179 | 0.05 | 0.06 | 0.379 | 73 | -0.01 | 0.04 | 0.842 | | 106 |
| | 3 | 0.00 | 0.03 | 0.974 | | 163 | 0.07 | 0.06 | 0.264 | 67 | -0.03 | 0.04 | 0.479 | | 96 |
| PhenoAge DNAmAA | 1 | 0.03 | 0.02 | 0.242 | | 179 | 0.00 | 0.04 | 0.982 | 73 | 0.05 | 0.03 | 0.133 | | 106 |
| | 2 | 0.04 | 0.02 | 0.153 | | 179 | 0.01 | 0.04 | 0.795 | 73 | 0.05 | 0.03 | 0.098 | | 106 |
| | 3 | 0.03 | 0.03 | 0.302 | | 163 | 0.00 | 0.04 | 0.945 | 67 | 0.02 | 0.03 | 0.474 | | 96 |
| GrimAge DNAmAA | 1 | -0.02 | 0.04 | 0.638 | | 179 | -0.10 | 0.08 | 0.222 | 73 | -0.01 | 0.05 | 0.816 | | 106 |
| | 2 | -0.03 | 0.04 | 0.477 | | 179 | -0.08 | 0.08 | 0.356 | 73 | -0.01 | 0.05 | 0.769 | | 106 |
| | 3 | -0.06 | 0.05 | 0.208 | | 163 | -0.14 | 0.09 | 0.152 | 67 | -0.04 | 0.06 | 0.473 | | 96 |

Note: DNAmAA = DNA methylation age acceleration, SE = standard error.

**Supplementary Table 7: Logistic regression of increase in DCSI of one or more points vs. no increase in DCSI (dichotomized) on 7-CpG DNAmA at baseline.** Model 1: no covariates; Model 2: DCSI (T0); Model 3: Model 2 + chronological age (T0) + sex (if applicable); Model 4: Model 3 + smoking (packyears, T0) + alcohol consumption (g/d, T0) + diabetes medication (yes/no, T0) + BMI (T0). Included are all participants with diagnosed diabetes mellitus at baseline.

| Model | Estimate | SE | OR | p-value | | n |
|---|---|---|---|---|---|---|
| Women and Men | | | | | | |
| 1 | 0.047 | 0.030 | 1.048 | 0.114 | | 101 |
| 2 | 0.046 | 0.030 | 1.047 | 0.127 | | 101 |
| 3 | 0.048 | 0.030 | 1.049 | 0.117 | | 101 |
| 4 | 0.040 | 0.033 | 1.041 | 0.219 | | 89 |
| Women | | | | | | |
| 1 | -0.040 | 0.051 | 0.961 | 0.429 | | 36 |
| 2 | -0.042 | 0.051 | 0.959 | 0.414 | | 36 |
| 3 | -0.037 | 0.052 | 0.964 | 0.477 | | 36 |
| 4 | -0.004 | 0.061 | 0.996 | 0.949 | | 33 |
| Men | | | | | | |
| 1 | 0.100 | 0.042 | 1.105 | 0.017 | * | 65 |
| 2 | 0.099 | 0.042 | 1.104 | 0.019 | * | 65 |
| 3 | 0.100 | 0.043 | 1.105 | 0.019 | * | 65 |
| 4 | 0.101 | 0.051 | 1.106 | 0.045 | * | 56 |

Note: T0 = baseline, SE = standard error, OR = odds ratio.

**Supplementary Table 8: Linear regression analysis of change in DCSI per year of follow-up time on 7-CpG DNAmAA at baseline.** Model 1: no covariates; Model 2: chronological age (years, T0); Model 3: Model 2 + sex (if applicable); Model 4: Model 3 + smoking (packyears, T0), Model 5: Model 4 + alcohol intake (g/day, T0); Model 6: Model 5 + antidiabetic medication; Model 7: Model 6 + BMI (T0).

| Model | | Estimate | SE | p-value | | n |
|---|---|---|---|---|---|---|
| Women and Men | | | | | | |
| | 1 | 0.006 | 0.003 | 0.080 | | 101 |
| | 2 | 0.006 | 0.003 | 0.082 | | 101 |
| | 3 | 0.006 | 0.003 | 0.076 | | 101 |
| | 4 | 0.006 | 0.003 | 0.086 | | 94 |
| | 5 | 0.006 | 0.003 | 0.094 | | 93 |
| | 6 | 0.006 | 0.003 | 0.090 | | 93 |
| | 7 | 0.005 | 0.003 | 0.189 | | 89 |
| Women | | | | | | |
| | 1 | -0.005 | 0.005 | 0.376 | | 36 |
| | 2 | -0.004 | 0.005 | 0.479 | | 36 |
| | 4 | -0.003 | 0.005 | 0.590 | | 35 |
| | 5 | -0.003 | 0.005 | 0.589 | | 34 |
| | 6 | -0.002 | 0.006 | 0.745 | | 34 |
| | 7 | -0.002 | 0.006 | 0.772 | | 33 |
| Men | | | | | | |
| | 1 | 0.011 | 0.004 | 0.007 | ** | 65 |
| | 2 | 0.011 | 0.004 | 0.008 | ** | 65 |
| | 4 | 0.011 | 0.004 | 0.012 | * | 59 |
| | 5 | 0.010 | 0.004 | 0.013 | * | 59 |
| | 6 | 0.010 | 0.004 | 0.014 | * | 59 |
| | 7 | 0.009 | 0.004 | 0.042 | * | 56 |

Note: SE = standard error.
